# Non-Metastatic Axillary Lymph Nodes Have Distinct Morphology and Immunophenotype in Obese Breast Cancer patients at Risk for Metastasis

**DOI:** 10.1101/2023.04.14.23288545

**Authors:** Qingyuan Song, Kristen E. Muller, Liesbeth M. Hondelink, Roberta M. diFlorio-Alexander, Margaret Karagas, Saeed Hassanpour

**Affiliations:** Department of Biomedical Data Science, Dartmouth College, 1 Medical Center Drive, HB 7261, Lebanon, NH, 03756, USA; Department of Pathology, Dartmouth-Hitchcock Medical Center, 1 Medical Center Drive, Lebanon, NH, 03756, USA; Department of Pathology, Leiden University Medical Center, Albinusdreef 2, 2333 ZA Leiden, The Netherlands; Department of Radiology, Dartmouth-Hitchcock Medical Center, Geisel School of Medicine at Dartmouth, Dartmouth Cancer Center, 1 Medical Center Drive, Lebanon, NH, 03756, USA; Department of Epidemiology, Dartmouth College, 1 Medical Center Drive, Lebanon, NH, 03756, USA; Department of Computer Science, Dartmouth College, Hanover, NH, 03755, USA

**Author notes:** Corresponding Author: Saeed Hassanpour, PhD, One Medical Center Drive, HB 7261, Lebanon, NH 03756. These authors contributed equally to this work (Qingyuan Song, Kristen E. Muller).

## Abstract

Obese patients have worse breast cancer outcomes than normal weight women including a 50% to 80% increased rate of axillary nodal metastasis. Recent studies have shown a potential link between increased lymph node adipose tissue and breast cancer nodal metastasis. Further investigation into potential mechanisms underlying this link may reveal potential prognostic utility of fat-enlarged lymph nodes in breast cancer patients. In this study, a deep learning framework was developed to identify morphological differences of non-metastatic axillary nodes between node-positive and node-negative obese breast cancer patients. Pathology review of the model-selected patches found an increase in the average size of adipocytes (p-value=0.004), an increased amount of white space between lymphocytes (p-value<0.0001), and an increased amount of red blood cells (p-value<0.001) in non-metastatic lymph nodes of node-positive breast cancer patients. Our downstream immunohistology (IHC) analysis showed a decrease of CD3 expression and increase of leptin expression in fat-replaced axillary lymph nodes in obese node-positive patients. In summary, our findings suggest a novel direction to further investigate the crosstalk between lymph node adiposity, lymphatic dysfunction, and breast cancer nodal metastases.

## Introduction

Breast cancer is the most prevalent cancer in women worldwide, and it is one of the leading causes of death in women^1,2^. Obesity, currently affecting over 30% adult females in the United States^3^, significantly increases the incidence and worsens the prognosis of breast cancer patients, among all breast cancer subtypes^4,5^. Specifically, obese women are 50-80% more likely to develop axillary metastasis^4–6^ and have higher breast-cancer specific mortality than normal weight women^7^.

The interaction between obesity, immunity, and breast cancer progression is complex, and the understanding of this link is an evolving field of research. Studies have demonstrated an increased risk of poor cancer prognosis among obese patients with ectopic fat within organs such as liver, muscle, and heart^8–10^. Enlarged adipocytes within ectopic fatty depots leads to the secretion of pro-inflammatory cytokines and metabolic dysregulation that creates a tumor-permissive micro-environment^6^. In particular, leptin has been identified as an adipokine that is increased in breast cancer patients^11,12^. The adipocyte-rich environment can also provide local fatty acids to fuel tumor growth^13,14^. With the prevalence of obesity rapidly increasing in almost all countries^15^, further investigation of the underlying mechanisms that put obese patients at an increased risk for axillary nodal metastases is crucial. Findings could support the evaluation of lymph node fat content in the workup of breast cancer patients and may inform prognosis, personalized treatment strategies and future targeted therapies.

As was recently shown by Almekinders et al, peritumoral breast hyperadiposity results in local steroid hormone biosynthesis and endocrine dysregulation, and is a potentially strong prognostic biomarker for invasive breast cancer in patients with ductal carcinoma in situ (DCIS)^16^. Prior studies have shown that non-metastatic axillary nodes may be enlarged by excess hilar fat deposition and are more commonly seen in women with obesity^17,18^. Fat-enlarged non-metastatic axillary lymph nodes identified on mammography and breast MRI are associated with a high risk of axillary metastasis in obese patients with invasive breast cancer, and this association is maintained when adjusting for patient and tumor characteristics^19^. Our hypothesize is that, hyperadiposity and micro-immune dysregulation may also occur in axillary nodal adiposity, creating a pre-metastatic niche for nodal metastases in women with invasive breast cancer. Our study aimed to investigate whether there are changes in the morphology and immunophenotype of nodal hyperadiposity and the immune microenvironment in non-metastatic axillary lymph nodes that are associated with lymph node metastasis in breast cancer.

## Methods

We developed and trained a deep learning (DL) framework to identify differences in morphological patterns in non-metastatic axillary lymph nodes from node-positive and node-negative patients. The DL model evaluated scanned whole slide images (WSI) of hematoxylin and eosin (H&E) stained slides using a histological image feature extractor to learn interpretable morphological patterns that may be shed light on underlying structural changes that contribute to the increased risk of nodal metastases in obese patients. We also accessed possible alterations of lymphatic, lipid and metabolic-related protein expression related to nodal metastasis through a detailed immunohistochemistry (IHC) analysis.

### Study Population and Data Collection

The study included obese patients (BMI > 30 kg/m^2^) with histologically confirmed invasive breast cancer who underwent sentinel lymph node excision (SLN) or axillary lymph node dissection (ALND) at Dartmouth-Hitchcock Medical Center (DHMC), Lebanon, NH. A total of 180 breast cancer cases were identified, including 88 patients with axillary nodal metastasis (labeled as node-positive) and 92 patients without nodal metastasis (labeled as node-negative). Patients’ demographics, clinical, and pathological information including age and BMI at the time of cancer diagnosis, tumor size, tumor grade, hormone receptor and HER2 status, and the presence of lymphovascular invasion (LVI) were collected from electronic medical records (**Table 1**). All patients included in this study did not receive neo-adjuvant treatment, including chemotherapy, prior to surgery. The detailed data collection workflow is illustrated in **Supplementary Figure 1**.

**Table 1.** Study cohort characteristics.

|  | Overall | Node-Negative | Node-positive |
| --- | --- | --- | --- |
| n | 180 | 92 | 88 |
| Age (years), mean (SD) | 60.9 (11.0) | 62.1 (9.9) | 59.6 (11.9) |
| BMI, mean (SD) | 36.4 (5.8) | 36.5 (6.1) | 36.3 (5.6) |
| Tumor Grade, n (%) |  |  |  |
| 1 | 26 (14.9) | 17 (19.5) | 9 (10.2) |
| 2 | 94 (53.7) | 43 (49.4) | 51 (58.0) |
| 3 | 55 (31.4) | 27 (31.0) | 28 (31.8) |
| Tumor Size, mean (SD) | 28.5 (23.6) | 24.1 (19.6) | 32.8 (26.4) |
| Molecular Subtypes, n (%) |  |  |  |
| ER+ and/or PR+, HER2- | 151 (84.8) | 74 (82.2) | 77 (87.5) |
| HER2+ | 11 (6.2) | 3 (3.3) | 8 (9.1) |
| TNBC | 16 (9.0) | 13 (14.4) | 3 (3.4) |
| LVI, n (%) |  |  |  |
| Absent | 96 (55.5) | 66 (76.7) | 30 (34.5) |
| Present | 77 (44.5) | 20 (23.3) | 57 (65.5) |

Abbreviations: SD – standard deviation; ER – estrogen receptor; PR – progesterone receptor; HER2 – human epidermal growth factor receptor 2; TNBC – triple-negative breast cancer; LVI – lymphovascular invasion.

### WSI of Axillary Lymph Nodes

Breast cancer patients underwent SLN or ALND at the time of their breast cancer surgery(partial or full mastectomy). Lymph nodes received from breast cancer patients were grossed according to standard procedures; all nodes were dissected longitudinally into 2 mm increments and submitted completely for histologic evaluation of metastatic disease. Tissue is formalin fixed and embedded into paraffin blocks (FFPE), sliced at 5-micron sections and stained with Hematoxylin and Eosin (H&E) for microscopic evaluation. The lymph node slides were retrieved and reviewed by a board-certified anatomic pathologist who specializes in breast pathology (KEM). For both study sets (node-positive and node-negative groups), only the non-metastatic lymph nodes were chosen for evaluation. Lymph nodes with metastatic foci of cancer were excluded from the dataset. The slides were scanned using the Leica Aperio-AT2 scanner (Leica Biosystem, IL, USA) at 20x and stored in SVS image format. A total of 636 WSI (303 node-positive and 333 node-negative) were collected.

### Lymph Node Annotation and Patch Extraction from WSI

The data preparation pipeline is illustrated in **Figure 1a**. We manually annotated regions of axillary lymph nodes containing lymphoid tissue and intranodal adipocytes on WSI with ASAP software (Version 1.9; Netherlands, 2018). Patches of 224×224 pixels were extracted from the pathologist-annotated regions at 10x magnification level to reduce the number of resulted patches. A total of 575,906 patches were extracted from the annotated regions from the H&E WSI (251,341 from node-positive patients and 324,565 from node-negative patients).

**Figure 1.**
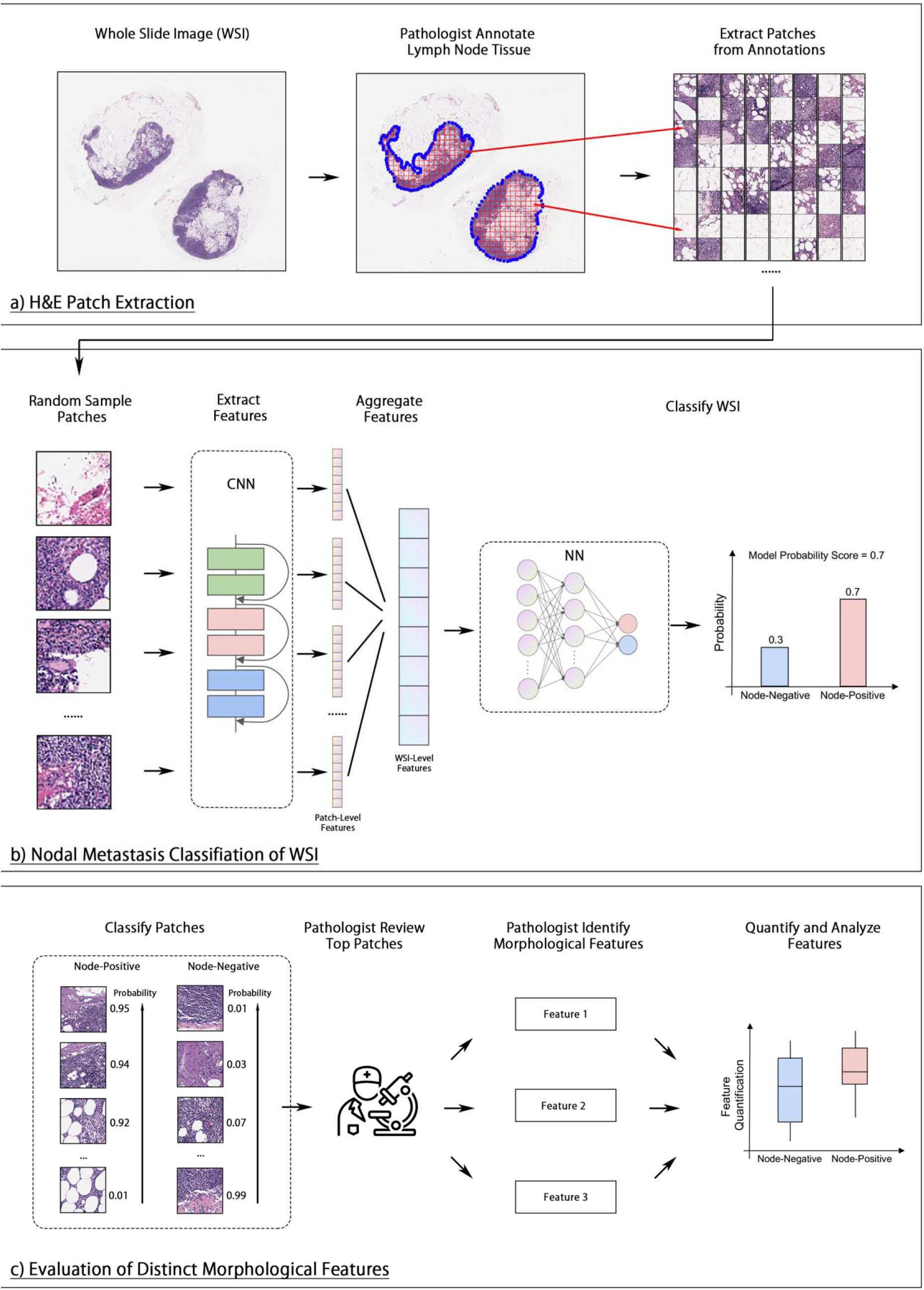
Summary of study design. a) Pathologist’s annotation of axillary lymph nodes (circled in blue) and patch extraction from WSI for use in model training. b) A CNN was used to extract image features from random sampled patches from a WSI. The resulted feature vectors were then averaged and fed into a fully connected NN to predict the node-positive probability of the WSI. c) The trained model was applied to each patch to classify their nodal status. Then, the patches were ranked by their model probability score within their label groups. The top-ranked patches were then examined by a pathologist to identify any differences in morphological features between the node-positive and -negative classes. These features were then quantified and evaluated for statistical differences.

### DL Framework for Histological Feature Extraction

We trained a DL framework based on previous work including Wulczyn et al.^20^ and Jiang et al.^21^ to classify the patients’ breast cancer nodal status using the non-metastatic lymph node histopathology. Given that the number of WSIs per patient varied, the patients’ nodal status was assigned to each WSI as the label, and the model was trained to classify each WSI separately. To start, randomly selected patches from each WSI were input into a convolutional neural network (CNN) with a ResNet-18 backbone and with ImageNet-pretrained weights^22,23^. The patches were converted to patch-level feature vectors of size 512, which were then averaged into a WSI-level feature vector. The resulting vector was fed into a 2-layer fully connected neural network (NN), with a hidden layer of 128 neurons and an output size of 2, indicating predictions of node-positive and negative classes. Lastly, a SoftMax normalization was applied to the model output to generate a probability score. The WSI was classified as node-positive if the resulting probability score was greater than 0.5, or as negative otherwise. The patient-level classification was determined by averaging the probability scores of all WSIs from the patient, and then converting to average score to a binary outcome using a threshold of 0.5. The model pipeline is illustrated in **Figure 1b**. The detailed model training and evaluation pipeline is shown in **Supplementary Materials I**.

### Model Visualization and Statistical Analysis of Morphological Features

**Figure 1c** illustrates the process of identifying morphological features from model-selected patches, as well as the quantification and statistical analysis of these features. Specifically, the trained model was applied to all patches from each patient to calculate a patch-level probability score. The 64 most-predictive patches from each patient were selected and grouped by patients’ labels for pathologist review. A pathologist (LH), blinded to the patients’ labels and nodal status, reviewed the selected patches to search for any distinct histological patterns between these two groups. The visual findings of morphological patterns were subsequently quantified using image processing techniques (**Supplementary Materials III**). The statistical significance of the difference in the quantified patterns between the two classes was determined by the Mann-Whitney U test, with a statistical significance considered at a p-value of less than 0.05.

### Immunohistochemistry (IHC) of Selected Markers

A total of 30 lymph node samples were selected for IHC analysis. Our goal was to select a combination of immunohistochemical stains to evaluate T-cell (CD3) and B-cell (CD20) proportions, fatty acid metabolism (fatty acid synthase (FASN), lipoprotein lipase (LPL), Spot14, and fatty acid translocase (CD36)), and adipose inflammation (leptin, adiponectin, tumor necrosis factor α (TNFα), and interleukin-6 (IL6)). A radiologist selected lymph node samples based on the extent of fat replacement within the lymph nodes using radiologic studies, with 15 samples obtained from patients who had positive lymph nodes and 15 from patients who had negative lymph nodes. Specifically, the 15 node-positive samples had high levels of fat replacement, while and 15 node-negative samples had low levels of fat replacement. Briefly, slides were deparaffinized and rehydrated. Primary antibodies against CD3 (Leica Cat# PA0553, Clone LN10), CD20 (Leica Cat# NCL-L-CD20-L26, Clone L26), FASN (Abcam Cat# ab22759, Polyclonal), LPL^24^, Spot14^25^, CD36 (Abcam Cat# ab133625, Clone EPR6573), leptin (Abcam Cat# ab16227, Polyclonal), adiponectin (Abcam Cat# ab75989, Clone EPR3217), TNF (Abcam Cat# ab220210, Clone TNFA/1172), IL6 (Abcam Cat# ab9324, Clone 1.2-2B11-2G10) were processed and stained according to manufacturers’ protocols. The expression level of the IHC stains were digitally quantified; the detailed methodology is described in **Supplementary Materials IV**.

## Results

### DL-Aided Interpretation of Distinct Lymph Node Morphological Features

Our DL-model trained on the WSI patches achieved an area under the receiver operating characteristic curve (AU-ROC) of 0.67 (95% CI: 0.59, 0.75) and the detailed model performance is shown in **Supplementary Table 2**. Through examination of the model-selected predictive patches, pathologists identified notable variations in histological characteristics between node-positive and node-negative cases. These differences were confirmed with our quantification metrics using our image analysis pipeline. The histological features that are considered and quantified in this study include: the number and size of adipocytes, the proportion of white space between the lymphocytes (defined by white space-lymphoid ratio), and the proportion of red blood cells (defined by erythrocyte-lymphoid ratio). The results and some examples of these measurements and analysis are illustrated in **Figure 2**, and further details on the quantification methodology can be found in the **Supplementary Materials III**. Based on this analysis, we found that node-positive patients had significantly larger intranodal adipocytes, with more variation in adipocyte size, compared to node-negative patients (**Figure 2a**). Additionally, there was a significantly increased proportion of white space between lymphocytes and red blood cells in the lymph node parenchyma of node-positive patients (**Figures 2b and 2c**). These morphological differences were also significantly distinct between the model-assigned labels (**Supplementary Figure 2**). In addition, we found a positive correlation between the increase of average size of adipocytes and higher white space-lymphoid ratio with a Pearson’s correlation coefficient of 0.26 (p-value < 0.001) (**Supplementary Figure 3**).

**Figure 2.**
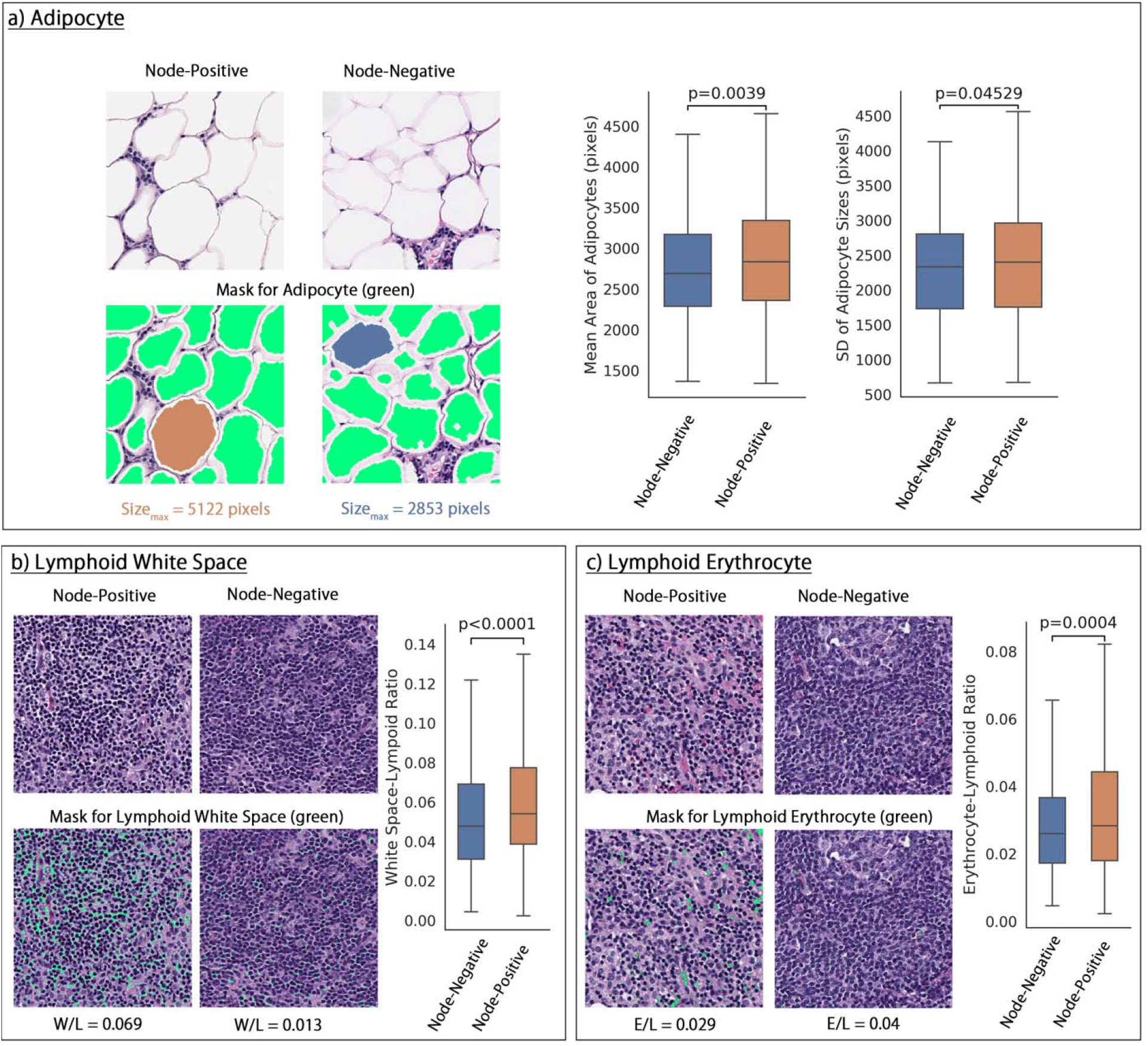
Distinct morphological characteristics of axillary lymph nodes in node-positive and node-negative patients. **a)** Left: example quantification of lymph node adipocyte size in fat-enlarged lymph nodes of node-positive and node-negative patients. The largest adipocyte from each patch was highlighted with orange or blue. Right: Box and whisker plot of the average size of adipocyte and the standard deviation (SD) of the adipocyte size in node-positive and - negative samples. **b)** Left: example quantification of lymphoid white space defined by white space-lymphoid ratio (W/L). Right: Box and whisker plot of the W/L ratio in node-positive and - negative samples. **c)** Left: example quantification of red blood cells in lymph node tissue defined by erythrocyte-lymphoid ratio (E/L). Right: Box and whisker plot of the E/L ratio in node-positive and -negative samples.

### Distinct Expression Pattern of IHC Stains in Lymph Node Lymphoid and Adipose Tissue

Although no statistical significance was observed in IHC analysis, the visual analysis of distribution of IHC stains revealed that fat-enlarged lymph nodes from node-positive patients displayed a lower ratio between CD3+ and CD20+ cells due to decreased CD3 expression (**Figure 3a-i**). Specifically, the intensity of CD3 and CD20 is significantly positively correlated in non-fatty lymph nodes from node-negative samples (Pearson correlation coefficient = 0.83, p-value < 0.001), while no significant correlation was found in fat-replaced lymph nodes from node-positive samples (Pearson correlation coefficient = 0.28, p-value = 0.30) (**Figure 3a-ii**). **Figure 3a-iii** and **3a-iv** show examples of a fat-replaced node from a node-positive patient with low CD3+/CD20+, and a non-fat-replaced node from a node-negative patient with high CD3+/CD20+.

**Figure 3.**
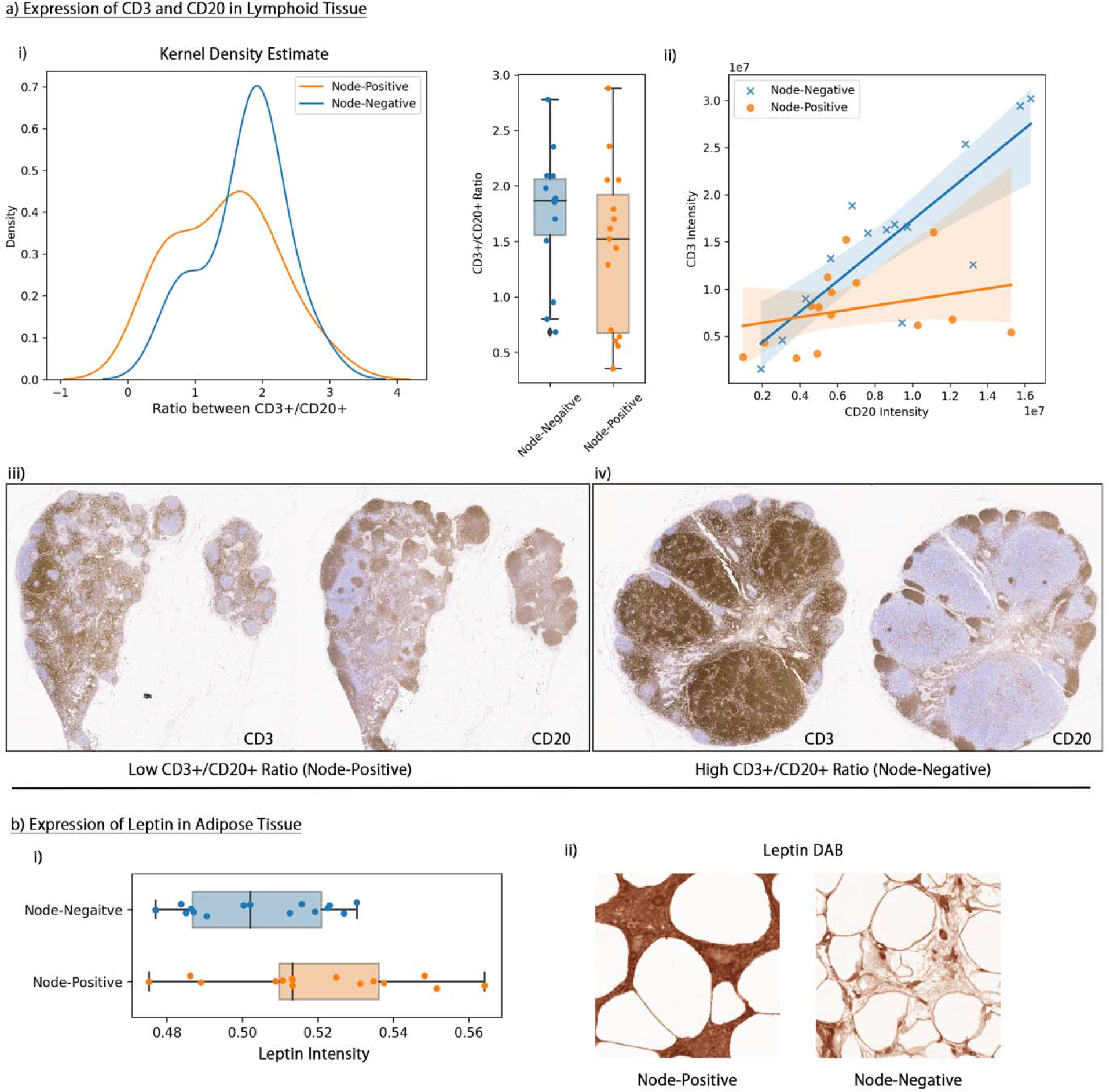
Distribution of IHC markers in node-positive and node-negative group. **a-i)** Kernel density estimate and boxplot of the distribution of CD3+/CD20+ ratio. **a-ii)** Correlation between CD3 and CD20 expression in node-positive and -negative node with fitted regression line. **a-iii)** A fat-enlarged node from a node-positive patient with low CD3+/CD20+ **a-iv)** A non-fat-replaced node from a node-negative patient with high CD3+/CD20+. **b-i)** Boxplot of distribution of leptin intensity in node-positive and -negative groups. **b-ii)** Comparison of leptin stain in the adipose tissue of lymph nodes from a node-positive and a node-negative sample.

Additionally, node-positive patients exhibited a slightly elevated expression of leptin in intranodal and perinodal adipose tissue compared to node-negative patients, as demonstrated in **Figure 3b**.

## Discussion

This study investigated the morphological and immunophenotypic characteristics of non-metastatic axillary lymph nodes in relation to the risk of breast cancer nodal metastasis among obese women. Using a cohort of 88 node-positive and 92 node-negative patients, we trained a DL model that identified several morphological features that were distinct in axillary lymph nodes from node-positive patients, including significantly increased average size of adipocytes, a higher proportion of white spaces within lymphoid tissue, and a higher number of erythrocytes within lymphoid tissue. In addition, the IHC analysis of 30 axillary lymph node samples showed visually decreased CD3 staining, lower CD3+/CD20+ ratios, and elevated leptin expression around nodal adipocytes in the fat-enlarged lymph nodes from node-positive patients, compared to that of the normal lymph nodes from node-negative patients. Together, our findings suggest a link between several histological characteristics of fat-enlarged axillary lymph nodes and nodal metastases in breast cancer patients with obesity.

Patients with obesity have poor breast cancer outcomes that are not fully explained by BMI. Understanding the factors that contribute to variable breast cancer outcomes among obese women is essential for improving prognosis and identifying potential therapeutic targets, ultimately leading to better outcomes for patients. Studies have found that adipose cells can contribute to cancer progression through the release of signaling molecules, extracellular proteins, lipids, and metabolites that support tumor growth and invasiveness^26^. Although there have many studies examining ectopic adipose located in and around organs, there is limited research investigating the impact of ectopic adipose within lymph nodes on cancer outcomes^27^. A recent study found that the presence of enlarged axillary lymph nodes due to fat infiltration as seen on mammography and breast MRI is correlated with an increased risk of nodal metastasis in obese patients when adjusting for patient and tumor characteristics^28^. Our current study identified distinct histologic features that could contribute to a tumor-promoting adipose microenvironment. These findings support the prior radiology study, which has shown a strong correlation between fat-enlarged lymph nodes and nodal metastases.

We observed a greater average adipocyte size within the axillary lymph nodes of node-positive patients than that of the node-negative patients. Previous research has established a link between adipocyte hypertrophy in obesity and impaired metabolic regulation that promotes cancer progression, partly due to hypoxia of hypertrophic adipocytes that leads to altered adipokine secretion and ensuing inflammation^29,30^. Our results align with those of Almekinders et al., who found a correlation between larger adipocyte size and an elevated risk of invasive breast cancer following DCIS. This underscores the importance of further research on the role of ectopic nodal adiposity in the development breast cancer nodal metastases.

Metastatic sites are selectively primed by the primary tumor even before the initiation of metastases occurs, resulting in a premetastatic niche that is devoid of cancer cells^31^. Our H&E analysis revealed an increased proportion of lymphoid white spaces in lymph nodes from node-positive patients, and a positive correlation between lymphoid white spaces and average adipocyte size of the lymph node. Possible theories to account for the increased white space include interstitial fluid caused by impaired lymphatic drainage^32^, which promotes migration of tumor cells. Another potential explanation is the production of soluble factors, such as protein ligands or extracellular matrix-modifying proteins that are secreted to induce remodeling of metastatic sites to facilitate seeding, as explained by Follain et al.^33^. Dissemination of tumor-related factors such as cytokines, chemokines, growth factors, matrix metalloproteinases, and extracellular vesicles have been shown to promote dissemination of tumor cells through stimulation of inflammatory cells and angiogenesis and leads to remodeling of the extracellular matrix^33–35^. We also noted an increased amount of extravasated red blood cells in the nodal tissue in the node-positive group. While speculation, one theory could be linked to the increased shear fluid forces in vessels with circulating tumor cells, while endothelial cell remodeling to promote tumor cell extravasation can provide an explanation for the leaky vasculature^36^. Vascular leakage and immunosuppression, among others, are changes that have been described in the premetastatic niche^31^. We postulate that the increased white space and extravasated red blood cells could represent morphologic evidence of preconditioning the premetastatic niche in axillary lymph nodes in obese patients.

Our downstream IHC analysis identified changes in immune cell populations and adipokine expression, though these results were not statistically significant due to the limited statistical power of the small sample size. CD3 intensity was lower in fat-enlarged nodes which may reflect decreased immune function in fatty nodes, a finding that has been reported in lymph nodes of obese mice^37,38^. Furthermore, non-metastatic fat-enlarged nodes of patients with nodal metastases also showed increased leptin expression. Leptin has been shown to stimulate proliferation, angiogenesis, migration, and metastasis in breast cancer cell lines by activating the oncogenic pathways, and high serum leptin is associated with a 2 to 5 fold increase in breast cancer risk^39^. Therefore, increased leptin expression in fat-enlarged nodes may also be associated with a pre-metastatic niche and requires further evaluation through large-scale IHC analysis.

This study is a pioneering effort that examined premetastatic morphological changes in lymph nodes in a cohort of obese patients with a focus on fat-enlarged lymph nodes. There are several limitations to our study. This study has a relatively small sample size, in particular the IHC analysis was restricted to a small number of patients due to limited resources. Our data is from a single institution which may introduce bias and chance of overfitting, and thereby limit the generalization of the findings to a larger population. A nested cross-validation approach was employed to mitigate this limitation. Further validation of our findings is necessary through large-scale, multi-center studies that include a diverse patient population and external validation. Our study is also limited by its retrospective design. Histologic images of axillary lymph nodes obtained via sentinel lymph node biopsy or axillary lymph node dissection were only available at the time of the cancer diagnosis. This temporal limitation makes it challenging to establish a causal relationship between the identified lymph node features and the development of nodal metastasis. However, future studies could investigate the relationship between lymph node characteristics related to nodal hyperadiposity and prospective outcomes, including cancer recurrence, distant metastasis, and long-term patient prognosis to further investigate the clinical importance of the identified morphological features.

In conclusion, this study identified several histopathological and immunohistochemical features of non-metastatic axillary lymph nodes in relation to breast cancer nodal metastases in obese breast cancer patients. These findings suggest that nodal hyperadiposity and alterations in the immune microenvironment may play a role in forming the pre-metastatic niche. These results emphasize the need for further research into increased lymph node adiposity. Further investigation into features of the fat-enlarged nodes that account for an increased likelihood of nodal metastases is warranted. If confirmed with larger studies, fatty axillary lymph nodes may serve as a prognostic imaging marker that can be readily assessed in all breast cancer patients. This information may inform personalized treatment strategies and targeted therapies in obese patients with breast cancer.

## Data Availability

All data produced in the present study are available upon reasonable request to the authors.

## Ethics Approval and Consent to Participate

This study, and the usage of human participant data in this project, were approved by the Dartmouth-Hitchcock Medical Center Institutional Review Board (IRB) with a waiver of informed consent.

## Author’s Contributions

All authors contributed to the design of the study. KEM was responsible for collecting the histological and immunohistochemistry image data, LMH annotated the histology images and RdA collected the clinical data. LMH and KEM analyzed and interpreted the histological findings. QS cleaned and preprocessed the data, implemented the methods, conducted statistical analysis, and wrote the first draft of the manuscript. All authors revised the manuscript and verified the presented results. SH supervised the study.

## Data Availability Statement

The datasets used and/or analyzed during the current study are available from the corresponding author on reasonable request.

