## Supplementary Materials for "Non-Metastatic Axillary Lymph Nodes Have Distinct Morphology and Immunophenotype in Obese Breast Cancer patients at Risk for Metastasis"

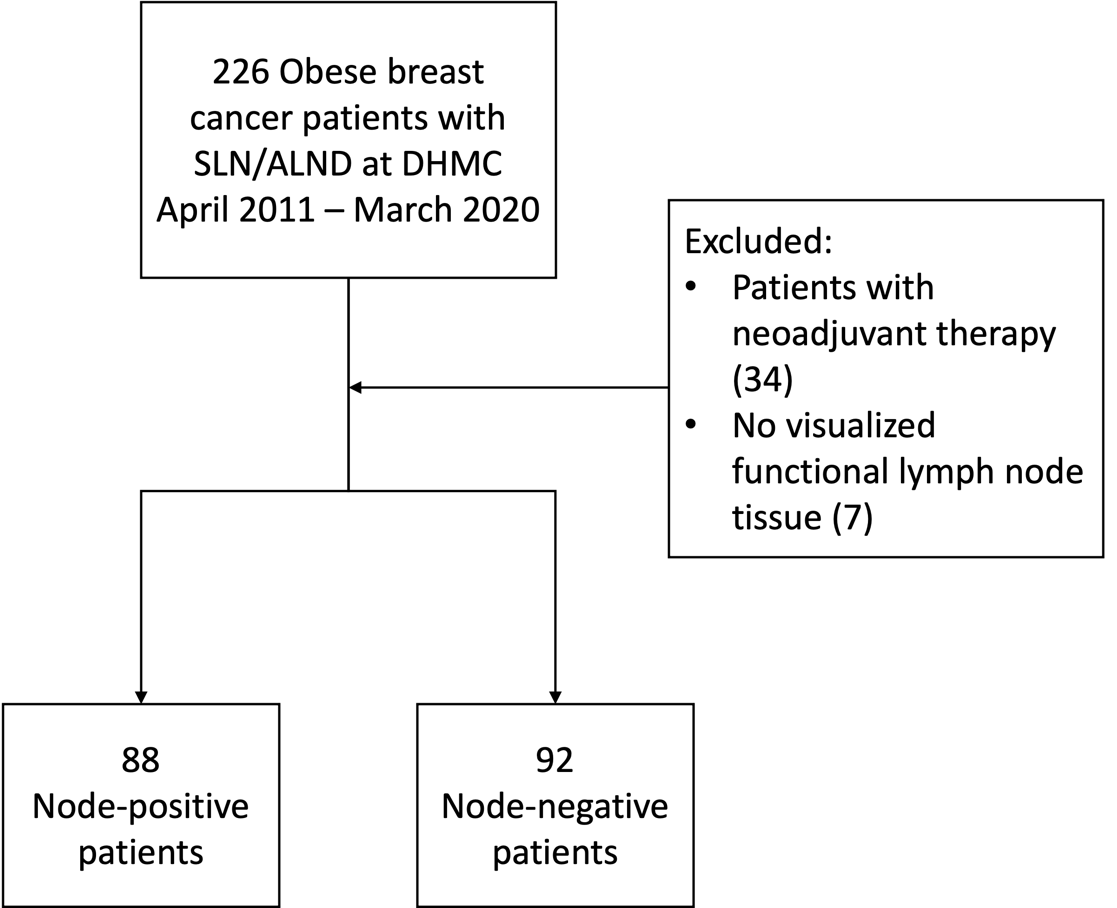

**Figure 1. Overview of Study Population Data Collection.**

### Supplementary Materials I: Model Training and Evaluation

Model Training and Evaluation

The model training and hyperparameter tuning was performed with 5-fold nested cross-validation stratified by patients’ nodal status. The dataset was split into 5 outer splits at the patient level to avoid information leak across different slides of the same patient. Each split was used as the held-out test once, while the rest of the patients were randomly split into 80% training set and 20% validation set for hyperparameter selection (inner loop). For each inner loop, we tuned the model hyperparameters including batch size, number of patches sampled during training, and initial learning rate for the backbone and the fully connected layer. The detailed hyperparameter configurations are shown in **Table 1**. The models were trained with an Adam optimizer and a cosine annealing learning rate scheduler for 300 epochs. The model with the lowest validation cross entropy loss during training was used for model evaluation. All models were trained using Nvidia Titan Xp graphic processing units with 12GB memory. The model was implemented and trained with Python version 3.81 and PyTorch version 1.132.

Image augmentation was applied to patches including random horizontal and vertical flip, random 90-degree rotation, and random color jittering during training. During cross-validation, for computational cost benefits, 500 patches were randomly chosen to select the optimal model (oversampled if the sample contains less than 500 patches). For evaluation, all patches were used to generate outer testing split predictions. The predictions of all outer test splits were aggregated and evaluated using overall accuracy, precision, recall, sensitivity, F1-score, and the area under the receiver operator characteristic curve (AU-ROC). The confidence intervals of the evaluation metrics were generated from 2,000 bootstrap samples.

**Table 1. Hyperparameters in the final model configuration**

|  | **Value** |
| --- | --- |
| **Training batch size** | 16 |
| **Start learning rate** |  |
| *Resnet-18 backbone* | $1\times{10}^{-6}$ |
| *Fully connected neural network* | $1\times{10}^{-4}$ |
| **Number of patches sampled per training iteration** | 32 |
| **Weight decay** |  |
| *Resnet-18 backbone* | 0.0001 |
| *Fully connected neural network* | $1\times{10}^{-5}$ |
| **Learning rate scheduler** | Cosine annealing with warm restart every 100 epochs |

### Supplementary Materials II: Detailed model performance

**Table 2. Slide and Patient-level DL Model Performance**

|  | **Accuracy** | **AU-ROC** | **F1** | **Precision** | **Recall** | **Specificity** |
| --- | --- | --- | --- | --- | --- | --- |
| **Slide-level** | 0.64  (0.60, 0.67) | 0.69  (0.65, 0.73) | 0.63  (0.59, 0.67) | 0.62  (0.56, 0.67) | 0.65  (0.59, 0.70) | 0.63  (0.58, 0.68) |
| **Patient-level** | 0.67  (0.60, 0.73) | 0.67  (0.59, 0.75) | 0.63  (0.54, 0.71) | 0.69  (0.58, 0.79) | 0.58  (0.48, 0.68) | 0.75  (0.66, 0.84) |

### Supplementary Material III: Quantification of Lymph Node Morphological Features

#### Detailed methods for quantification

##### Adipocyte

The adipocyte quantification method in this study is similar to that described in Osman et al.^3^. The image was transformed into the HSV format. A binary threshold of 5 was applied to the saturation channel of the image to eliminate background noise. An opening operation using a disk-shaped kernel was performed on the image, followed by a dilation operation using the same kernel. Finally, an opening operator using a disk-shaped kernel was applied to the inverse of the previous image to generate the mask for adipocytes.

The contour-finding algorithm was applied to the adipocyte mask to identify the area of individual adipocytes. Areas that were less than 500 pixels (i.e., small tissue gaps) or greater than 20000 pixels (i.e., background, large tissue gaps) were removed from the adipocyte areas.

##### Lymphoid White Space

The image was converted to HSV format. A binary threshold of 30 was applied to the saturation channel of the image to eliminate background noise and create a binary tissue mask. The tissue mask was then inverted to generate a mask for all white spaces.

The edges of the white space mask were filled in to eliminate intensities caused by the background. All connected components (objects) that had more than 200 pixels were removed from the white space mask. This step eliminated white spaces resulting from adipocytes and tissue gaps caused by broken tissues during laboratory procedures. The remaining mask is referred to as the lymphoid white space.

##### Lymphoid Erythrocyte

The image was transformed into the HSV format. Pixels within the intensity range of 0 to 10 and 160 to 180 (red color) from the hue channel were considered as erythrocytes.

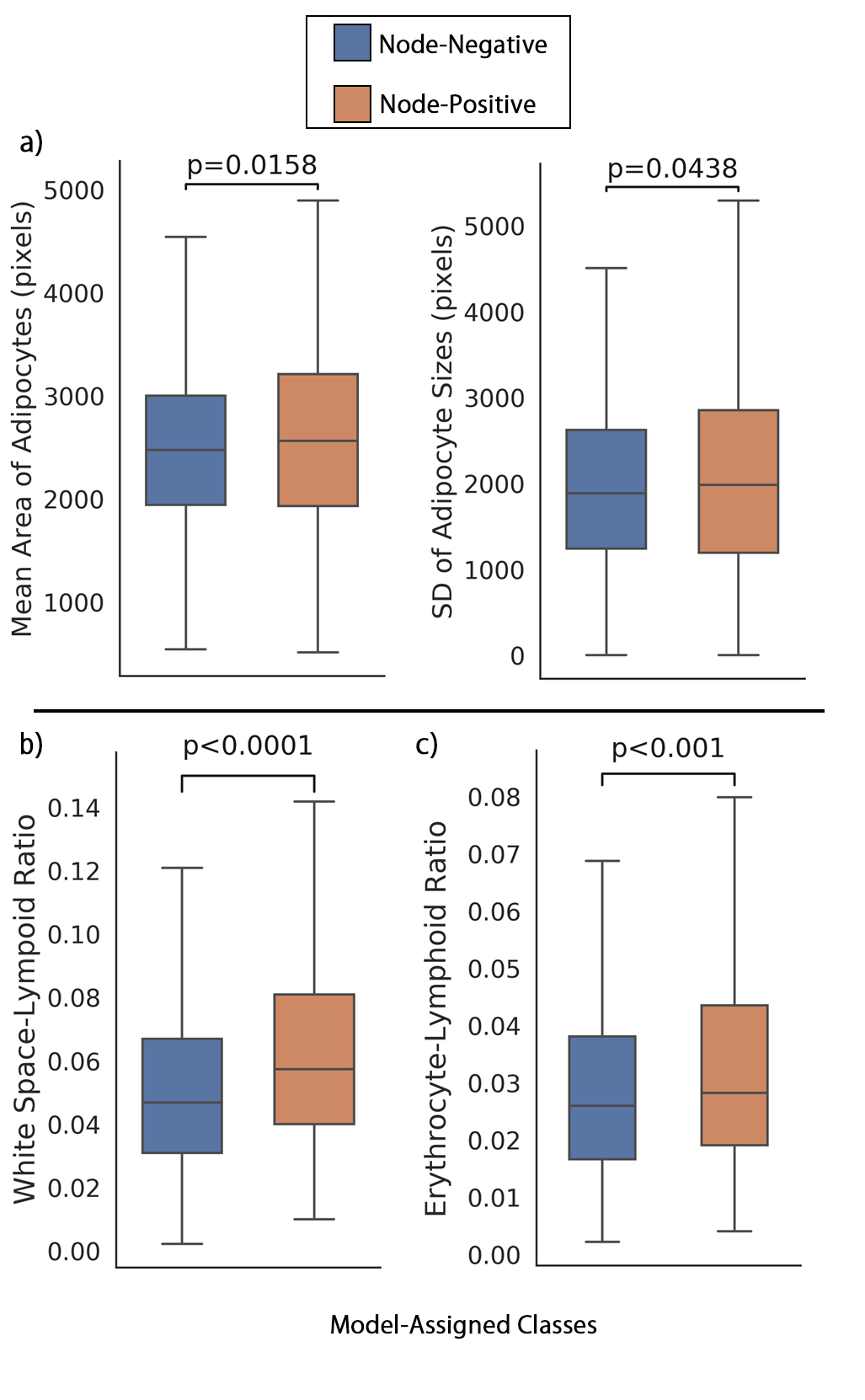

**Figure 2. Distribution of axillary lymph node morphological features in the model predicted class with p-value from Mann-Whitney U test.** a) Mean adipocyte size and standard deviation (SD) of adipocyte size was higher in model-assigned node-positive class. b) The lymph nodes from model-assigned node-positive class had higher white space-lymphoid ratio. c) The lymph nodes from model-assigned node-negative class had higher erythrocyte-lymphoid ratio.

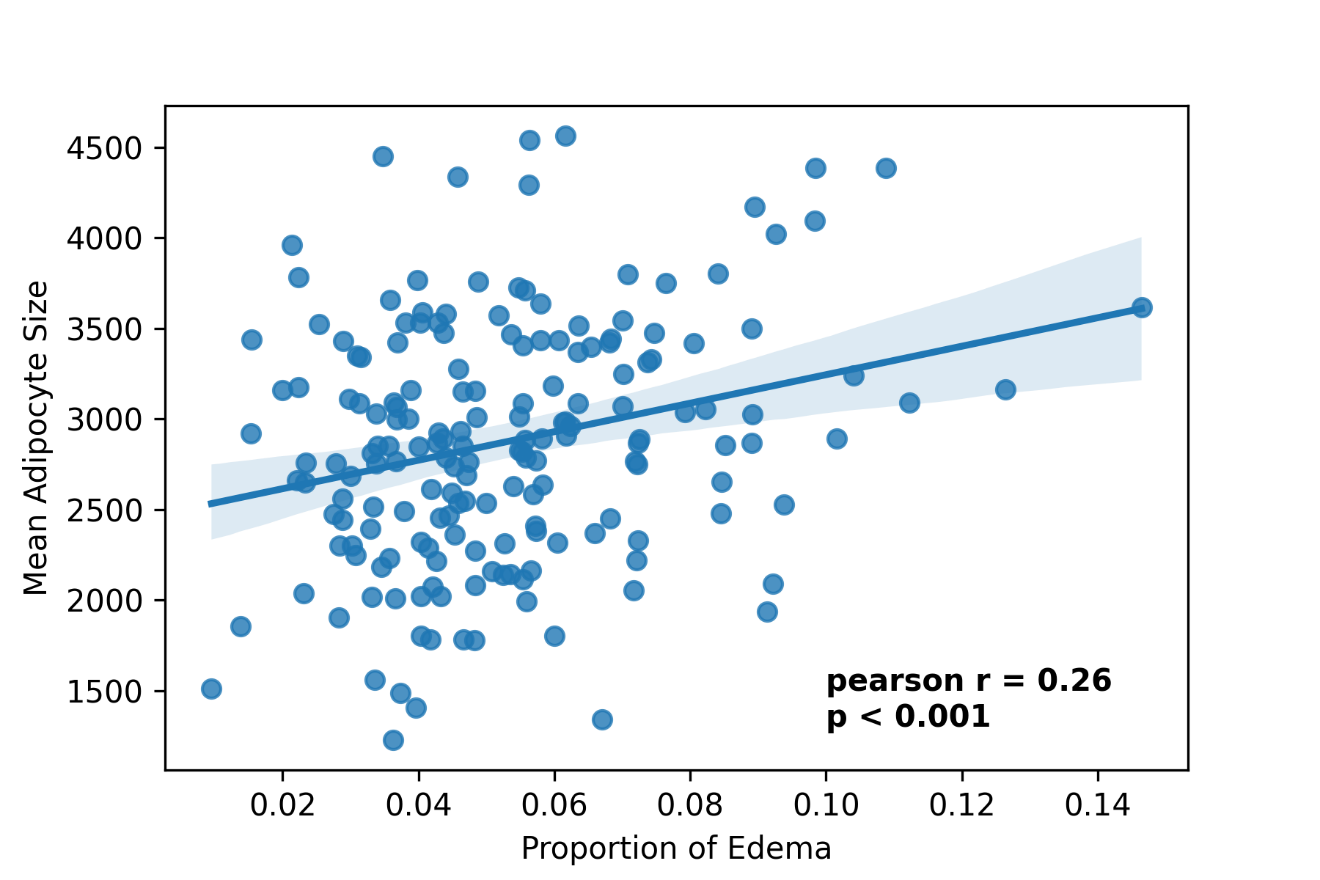

**Figure 3. Correlation between white space-lymphoid ratio and mean adipocyte size.** The white space-lymphoid ratio and mean adipocyte size of axillary lymph nodes are positively correlated with a Pearson correlation coefficient of 0.26 (p-value < 0.001). Blue line: fitted regression line. Light blue shade: 95% confidence interval of the fitted regression line.

### Supplementary Materials IV: Quantification of Immunohistochemical Stains (IHC)

IHC stains were quantified in the lymphoid tissue region, adipose tissue region, or both based on the location of expression of each stain.

From the whole slide image of the stained lymph node, all patches from areas containing stained tissue were extracted. Each patch was classified as adipose-rich or lymphocyte-rich using the adipocyte detector described in **Supplementary Material III**. The hematoxylin channel was separated from the patches using the Scikit-image package (version 0.19.2) with methods outlined in Ruifrok et al.^4^
